# Computed Tomography Radiomics Signatures: Sensitive biomarkers for clinical decision support in pancreatic cancer- a pilot study

**DOI:** 10.1101/2021.12.03.21267217

**Authors:** Abbas Habibalahi, Daniel Moses, Jared Campbell, Saabah Mahbub, Andrew P Barbour, Jaswinder S Samra, Koroush S Haghighi, Val J Gebski, David Goldstein, Ewa Goldys

**Affiliations:** ARC Centre of Excellence Centre for Nanoscale Biophotonics, University of New South Wales, Sydney, 2052, NSW, Australia; The Graduate School of Biomedical Engineering, University of New South Wales, Sydney, 2052, NSW, Australia; Department of Medical Imaging, Randwick Campus Hospitals, Sydney, 2052, NSW, Australia; The University of Queensland Diamantina Institute, The University of Queensland, Brisbane, 4102, Queensland, Australia; Queensland Melanoma Project, Princess Alexandra Hospital, Brisbane, 4102, Queensland, Australia; The University of Sydney, Northern Clinical School, Sydney, 2006, NSW, Australia; Prince of Wales Hospital, Prince of Wales Clinical School, University of New South Wales, 2052, NSW, Australia; NHMRC Clinical Trials Centre, University of Sydney, Sydney, 2006, NSW, Australia; Department of Medical Oncology, Prince of Wales Hospital, Sydney, 2052, NSW, Australia

## Abstract

**Aim:** To evaluate if suitably designed computed tomography (CT) radiomic signatures are sensitive to tumour transformation, and able to predict disease free survival (DFS) and overall survival (OS) time in patients with pancreatic cancer.

**Method:** Ethical approval by UNSW review board was obtained for this retrospective analysis. This study consisted of 27 patients with pancreatic cancer. Unsupervised principal component analysis was employed to evaluate the sensitivity of radiomic signatures to cancer presence and treatment. Further, optimised radiomic signatures were discovered using swarm intelligence and assessed for their capability to predict DFS and OS based on Kaplan-Meier analysis and receiver-operator characteristics (ROC) curves.

**Results:** We found that appropriate two radiomic signature are sensitive to cancer presence (area under the curve, AUC=0.95) and cancer treatment, respectively. Two other optimized radiomics signatures showed significant correlations with DFS and OS, respectively (p<0.05).

**Conclusion:** The CT radiomics signatures are an independent biomarkers which are modified when cancer is present and can help to estimate DFS and OS in patients. These signatures have the potential to be used to support clinical decision-making in pancreatic cancer treatment.

## 1. Introduction

Pancreatic cancer (PC), the 4th leading cause of cancer deaths, has a poor 5-year survival (<9%) while 430,000 people worldwide currently die from PC every year [1]. Multiple large-scale Phase III studies over the past two decades have produced relatively modest survival gains [2]. Patients with clearly resectable tumours comprise 15-20% of the presenting cohort at diagnosis. Surgical resection followed by multiagent adjuvant systemic therapy remains the number one treatment for those patients leading to median overall survival rates of 28-54 months[3]. An additional 20% is comprised of patients with either a borderline respectable tumour i.e. one with a high risk of incomplete resection [4] and those with locally advanced unresectable disease without systemic spread [5]. Retrospective cohort studies [6] and small prospective trials have suggested that neoadjuvant therapy presents an opportunity to increase the resectable cohort [3, 7].

Currently, local and distant recurrences following resection are frequent [8] and, for the 80% of patients with non-metastatic disease who have borderline resectable or locally advanced pancreatic cancer, 5-year overall survival is just 12% [9]. Biomarkers, including circulating tumor antigen CA19-9 and genomic markers, have not been prognostically accurate enough to identify patients who are most likely to benefit from surgery and/or additional interventions such as chemotherapy and radiotherapy [10].

Radiomics is an emerging quantitative approach to standard-of-care clinical images where automatically extracted quantitative image information is utilised to derive medically relevant conclusions [11]. The images are first inspected to identify a region of interest (ROI) which may comprise the whole tumor, specific areas, relevant organs or other points of reference. Boundaries of the ROI are “segmented by an operator or software and quantitative features are extracted, including both conventional, visually detectable differences in shape, intensity, or textures [12] as well as image differences which are too subtle to be perceived by human operators. These subtle differences are instead captured by a careful mathematical or data-driven analysis of the spatial distribution of image pixel intensities. In this way, the Radiomics approach overcomes the subjective nature of clinical image assessment [13]. This is important, in light of high human error rates, e.g. for assessing resectability the literature cites a 23% error rates [14] despite new international consensus definitions. Errors in image analysis are due to variable background and training of observers; thus, there is a significant need for a more objective methodology to derive clinical conclusions [15].

In this study, we developed and assessed several (N=4) radiomic signatures from CT images of pancreatic cancers potentially useful for different medical decision-making applications. A specific radiomic signature was developed and its sensitivity was evaluated to the presence of cancer. Further, a linear classifier was developed for potential automatic cancer segmentation. Next, a distinctive radiomic signature was constructed which is sensitive to the effect of chemotherapy and tumor progression based on base-line and pre-operative CT images. Finally, two separate radiomic signatures developed to predict outcome for both disease free survival (DFS) and overall survival (OS) using regression analysis combined with swarm intelligence[16]. These radiomic signatures were subsequently used to identify patient subpopulations with long or short survival time (disease free survival and overall) with performance assessed by Kaplan Meyer survival and receiver operator characteristic (ROC) analysis.

## 2. Materials and methods

### 2.1 Patient recruitment

GAP was a single-arm, multicentre, Phase II trial of perioperative chemotherapy utilizing nab-gem in patients with radiologically defined respectable PDAC [17]. Eligible patients were recruited from eight sites across Australia between 19 June 2012 and 30 June 2014[18]. Inclusion criteria were: aged 18 or older, ECOG performance status of 0–2 with confirmed PDAC, and resectable disease on CT/MRI based on established guidelines[19, 20]. This was defined as no evidence of any of extra-pancreatic disease, tumour abutment of the superior mesenteric artery (SMA) or coeliac axis (T4), portal vein (PV) infiltration of more than 180° of the circumference or occlusion of the superior mesenteric vein (SMV) or SMV-PV confluence. Eligibility was assessed at local institutional tumor boards. Major exclusions included borderline resectable tumour or locally advanced disease prior to registration,[20] previous radiotherapy to the upper abdomen, and significant medical conditions that prevented treatment.

All participants provided written informed consent. The protocol was centrally approved by the Sydney Local Health District Ethics Review Committee, Royal Prince Alfred Hospital. The Australasian Gastro-intestinal Trials Group (AGITG) was the study sponsor and it was coordinated by the National Health and Medical Research Council Clinical Trials Centre (NHMRC-CTC) at the University of Sydney. Specialised Therapeutics Australia supplied the nab-paclitaxel and provided untied financial assistance to conduct the trial. The trial was registered with the Australian and New Zealand Clinical Trial Registry (ACTRN12611000848909). Forty-two patients were enrolled in the trial, and where imaged using either multiphase CT, PET/CT or MRI. Portal venous phase CT images were chosen for analysis as they are the most consistent post contrast phase in terms of intravenous contrast concentration and rate of change. Twenty-seven of the patients had baseline CT studies which included a portal venous scan (Patient characteristics and exclusion reasons are detailed in supplementary material table 1, Consort flow diagram is detailed in supplementary figure 1).

### 2.2. CT imaging (baseline and pre-opp)

Baseline assessments were made in the 28 days prior to registration. Safety investigations were done at baseline and prior to each treatment cycle. A baseline three-phase CT scan of the chest/abdomen/pelvis or MRI-determined resectability and was repeated prior to surgery to assess response and resectability, and again after the second and fourth cycles of AT. 18FDG-PET combined with a low-dose CT scan was performed prior to Day 1 and on Day 15 (±2 days) of the first cycle of chemotherapy (CHT) to assess early metabolic response.

### 2.3. Segmentation and feature extraction

Image segmentation and radiomic analysis was performed using 3D Slicer [21] on portal venous phase images. Double blinded hepatobiliary radiologist segmented the tumour and a region of normal pancreatic tissue to produce 3D regions of interest (ROI). Radiomic features were generated using the Slicer Radiomic extension, Pyradiomics [22]. In this study different classes of quantitative imaging features, each describing a different property of a region of interest (ROI), were extracted. This features includes ROI shape features such as tumour size, diameter and circularity as well as heterogeneity [23]. Commonly used first-order (e.g., mean intensity) and second-order features (e.g., contrast and homogeneity) were captured [24]. This feature bank captures comprehensive information about the tumour, Radiomic feature dimensionality reduction and classification.

### 2.4. Data analysis

We developed 4 distinctive radiomic signatures (1) to detect tumour (tumour radiomic signature), (2) to assess treatment (treatment radiomic signature), (3) to predict outcomes based on DFS (DFS radiomic signature), and (4) to predict outcome based on OS (OS radiomic signature).

#### 2.4.1 Construction of tumour and treatment radiomic signatures

Tumour and treatment radiomic signatures were constructed by applying principal component analysis (PCA) to significant radiomic features (p<0.05) identified by Mann-Whitney U test (MWU) [25]. PCA is a standard unsupervised dimension reduction approach which is secure from overfitting. PCA projects the radiomic vectors onto the eigenvectors of the covariance matrix to elucidate useful variations of uncorrelated data. Top-ranked PCA scores capture the most significant portion of data variability (>90%) [26]. In this work, we simply considered top 2 ranked PCA scores obtained from tumor/normal image sections and baseline/pre-opp imaging as a 2D radiomic signature for tumour and treatment radiomic signatures, respectively.

#### 2.4.2 Construction of DFS and OS radiomic signatures

The DFS and OS radiomic signature need to be one dimensional to be able to predict the survival outcomes. Swarm intelligence and regression analysis were used simultaneously to select a limited number (N=5) of most useful prognostic features and construct from them the DFS and OS radiomic signatures [16]. Briefly, swarm intelligence replicates the progression of naive information-managing cooperating agents in a group aiming to reach a target [27]. In this work, the agents were radiomic feature subsets[16]. These agents repetitively evolve according to a pre-set evolution rule[28] to achieve the target. In our case the target was to maximise the goodness of fit (measured by the coefficient R^2^) of the linear regression curve to the patient feature data. R^2^ varies between 0 to 1 corresponding to the lowest to the best goodness of the fit, respectively. Initially, a number of agents (K=100 in this work) with random subsets of 5 feature are selected, and their R2 values are calculated. Next, these agents are iteratively updated until R^2^ converges to a highest value. One of such agents (feature subsets) whose R^2^ is close enough to that highest value is identified, and the coefficients of its linear fit are then recorded. At this point we have found an optimised linear combination of 5 features which represents the best fit to patients’ data, meaning that an optimum radiomic signature is constructed.

#### 2.4.3. Assessing radiomic signature based on cut-point optimization

DFS and OS radiomic signatures were used to divide patients into high and low risk outcomes based on cut point optimization and Kaplan-Meier (KM) analysis.

To identify patient subpopulations (long time survival vs. short time survival) using radiomic signature, a cut-point is required. To this end, a standard method called “X-tile” was employed as detailed in [29]. X-tile demonstrates the presence of substantial patient subpopulations and confirms the robustness of the relationship between radiomic signature and outcome by evaluating every possible subpopulation and optimizing cut-off point [29]. To evaluate the robustness of radiomic signature and corresponding cut-point to classify patients, Kaplan-Meier (KM) strategy was used as discussed in [30]. KM measures the fraction of subjects living for a certain amount of time after treatment. The treatment effect is assessed by the number of patients survived or saved after that intervention over a time period. The time commences from baseline imaging to the occurrence of death.

## 3. Results

In this study, we analysed 27 patents (current cohort) due to exclusion of 15 patients from our original patient cohort (N=42). Reasons for exclusion were, in the majority, technical, with 11 being due to the lack of cortal venous phase CT images for assessment (section 2.1) due to other scanning modalities being utilised (supplementary table 1). A demographic table (Table 1) details clinical variables and differences between the original GAP and current study cohort. There were only negligible differences between two cohort’s clinical variables mean values amounting to 1%, 1%, 7% and 3% for age, BMI, CEA and Ca19-9, respectively.

**Table 1.** Demographic table

| Continuous covariates | N | Mean | STD | MIN | Median | Max |
| --- | --- | --- | --- | --- | --- | --- |
| age | 27 | 62.92 | 9.68 | 42.8 | 64.5 | 78.64 |
|  | 42 | 64.1 | 8.66 | 42.8 | 65.3 | 78.6 |
| BMI | 27 | 26.47 | 5.2 | 19.03 | 25.45 | 37.1 |
|  | 42 | 27.1 | 5.77 | 18.5 | 26.1 | 47.7 |
| CEA | 27 | 5.9 | 8.6 | 0.5 | 3 | 32 |
|  | 42 | 5.44 | 7.93 | 0.5 | 2.7 | 32 |
| Ca19-9 | 27 | 731.55 | 1628.13 | 1 | 80 | 7305 |
|  | 42 | 703 | 1554 | 1 | 80.5 | 7305 |

### 3.1 Tumor radiomic signature

Figure 1 (a) represents radiomic signature values obtained from tumour regions (red points) and regions of normal pancreatic parenchyma (blue points) as detailed in Section 2.4.1 and Supplementary Material Table 2. It shows that tumour regions have distinctive radiomic signature (details in supplementary material table 2) from normal regions by forming a separate cluster (P<0.05). Further, radiomic signature (top 2 PCA scores) were fed to a linear classifier to predict data points labels. To approximate the sampling error due to a relatively small data set, a bootstrapping approach was used due to fairly small sample size [31]. To this end, data points were randomly resampled 100 times with the substitution from the original set of observations and then the corresponding ROC curves were obtained, and 95% confidence interval calculated. The performance of the Classifier was evaluated based on the ROC curve (Figure 1 (b)) and it was found to be very accurate (AUC=0.95±0.04).

**Figure 1.**
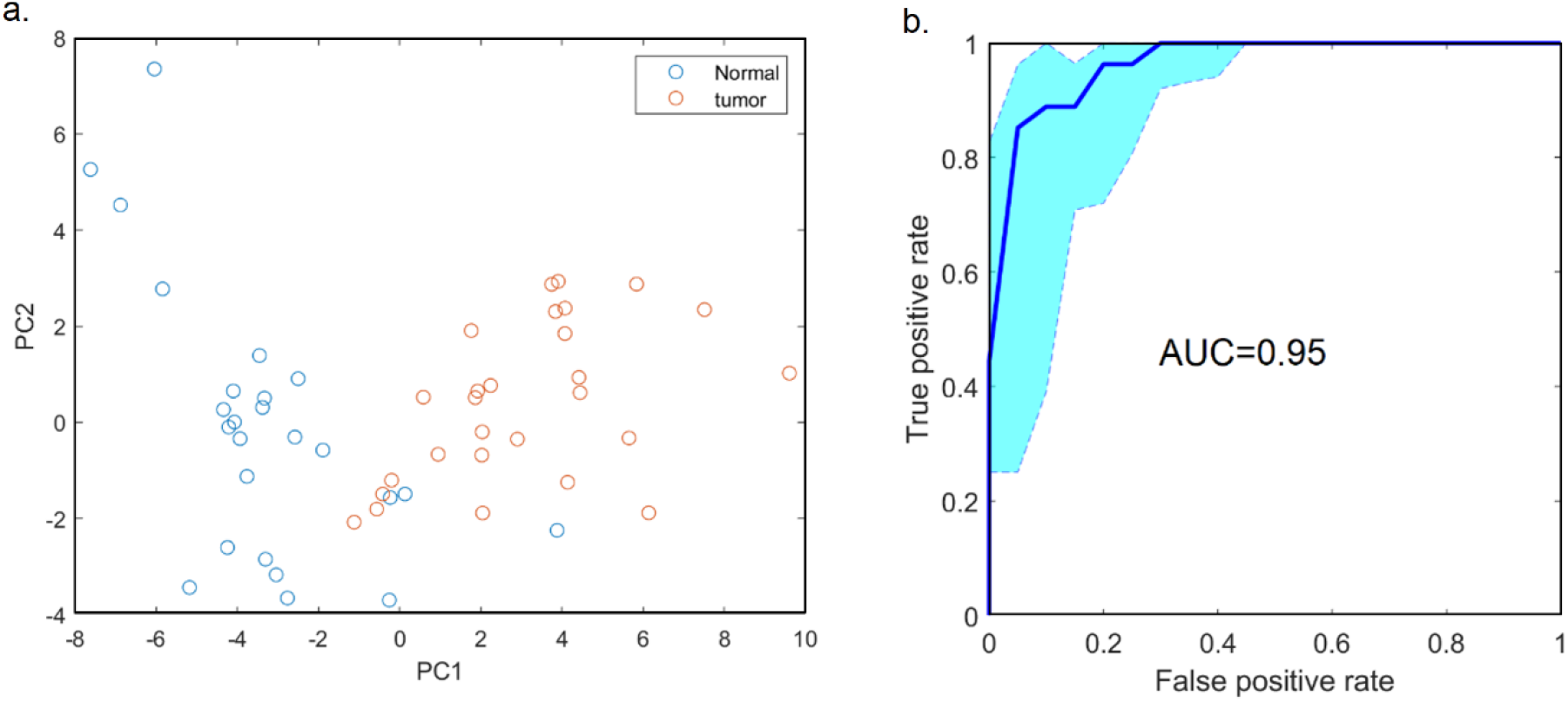
Tumour and normal pancreas radiomic signature. (a) PCA discriminative analysed (p<0.001 based on Mann-Whitney U-test applied on PC1) (b) ROC curve (AUC=0.95).

### 3.2. Treatment radiomic signature

We analysed radiomic signature for patients with both baseline and pre opp CT images (N=19). Figure 2 (a) shows radiomic signature (details in Section 2.4.1. Supplementary Material Table 3) obtained from pre-operative imaging data points (blue) and baseline images data points (red). Baseline datapoints form a cluster cantered away from the pre-op data points. Figure 2 (b) is the boxplots corresponding to the PC1 with significant difference (P<0.005). Visual assessment of Figure 2 (a) and (b) shows that 7 pre-opp data points are mixed with baseline clusters representing 82% patients radiomic signature modifies due to treatment and/or cancer development.

**Figure 2.**
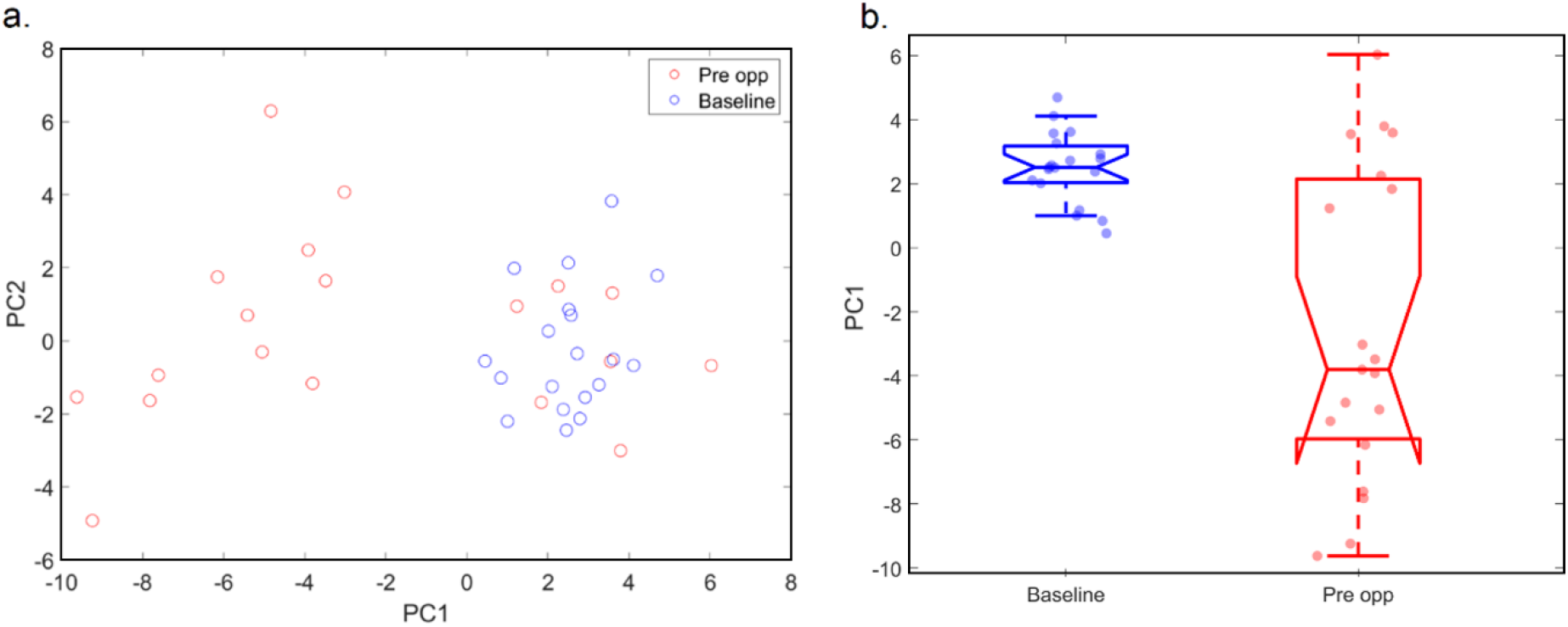
(a)Pre-op radiomic signature modification due to cancer development/ treatment. (b) Boxplots corresponding to the PC1 representing significant difference between baseline and preopp radiomic signature based on Mann-Whitney U-test (P<0.005).

**Figure 3.**
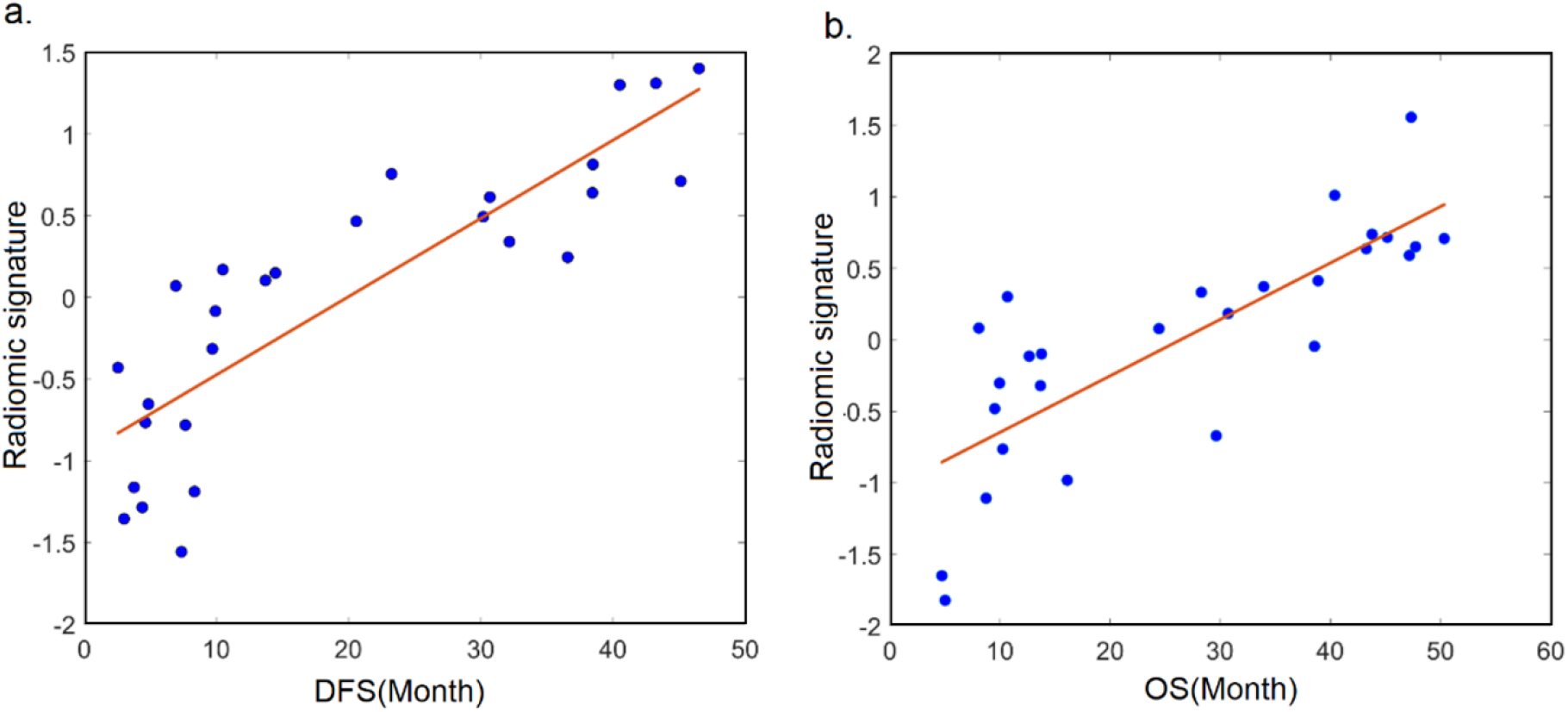
Radiomic signature to predict survival time. (a) DFS radiomic signature (b) OS radiomic signature

### 3.3. DFS and OS radiomic signatures

DFS and OS radiomic signatures (see Section 2.4.2 and Sup Mat Table 4 and 5) were discovered based on radiomic features in the baseline scans employing a limited number of features (N=5) to minimize overfitting risk. The goodness of OS and DFS radiomic was evaluated based on coefficient of determination (R^2^)[32] which was found to be 0.73 and 0.63 for DFS and OS radiomic signature, respectively.

### 3.4. DFS and OS radiomic signature classify patient with high and low risk outcomes

The X-tile method was applied to the DFS and OS radiomic signature obtained in section 3.3 to identify radiomic signature cutting point and dividing patient into two groups with high and low radiomic signature values. The values of the DFS and OS cutting points were found to be -0.4 and -0.1. Patient survival time with low and high radiomic signature values were compared against survival function from lifetime data using KM graphs. Figure 4 (a,b) shows the percentage survival of patient with low radiomic signature values (blue line) versus patients with high values (grey line) and demonstrate that both DFS and OS radiomic signatures could significantly (P<0.01) classify patients with long survival time from patients from short survival time. The classification performance (AUC) was found to be 0.94±0.07 and 0.87±0.17 for DFS and OS, respectively (details in supplementary materials figure 2).

**Figure 4.**
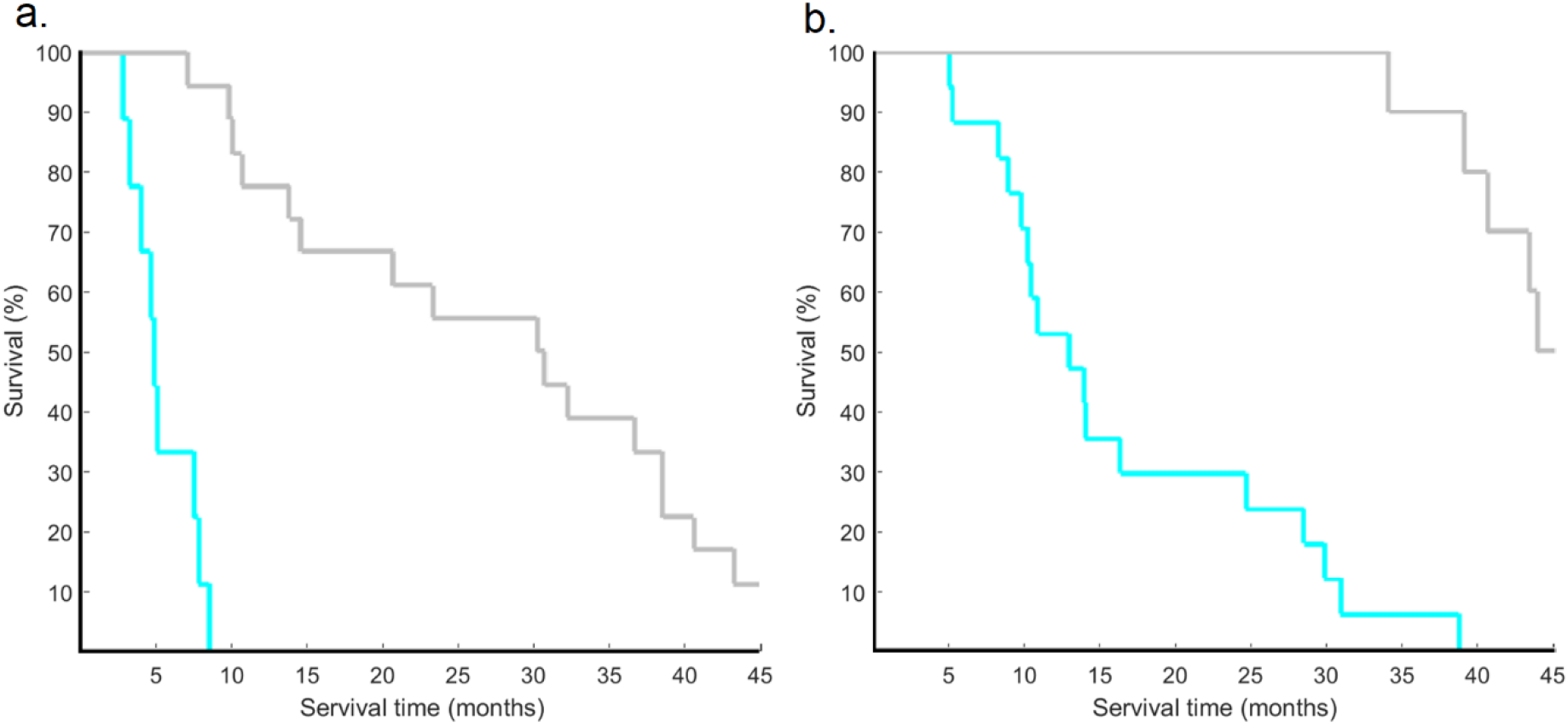
Kaplan- Meier analysis to evaluate survival time (a) Kaplan- Meier graph based on DFS (b) Kaplan- Meier graph based on OS.

## 4. Discussion

The rapid development of new medicinal options in the pancreatic cancer treatment requires biomarker discoveries for a reliable and correct pre-therapeutic patient classification and treatment assessment. In this communication, we demonstrate that radiomic analysis of pancreatic tumour paired with artificial intelligence has the ability to identify tumour sections and is sensitive to chemotherapy. We then demonstrate evidence for the functionality of radiomics signature to predict the survival rate in patients.

Pancreas cancer has poor outcomes and as treatment choices evolve there is a need to improve the prediction of outcomes and reduce inappropriate treatments unlikely to benefit patients. Recent data has shown the potential benefits of neoadjuvant therapy in improving survival compared to surgery alone [33]. The challenge remains to identify reliably those patients for whom this approach provides sufficient gain to justify the time and risk of side effects that are involved. In this communication, we demonstrate that radiomic analysis of pancreatic tumour paired with artificial intelligence has the ability to distinguish tumour from surrounding tissue tumour sections and demonstrates change with treatment exposure. We then demonstrate evidence for the functionality of baseline radiomics signature to predict the survival rate in patients.

Among the features tested in our study, zone entropy, demonstrating heterogeneity of an image, was common to the predictive model of radiomic signatures found for each of the tumour section classification, change with treatment and outcome response associated with DFS and OS. Our data conforms regarding entropy with the reports from meta-analyses of different tumour entities, across imaging modalities as a promising candidate feature to evaluate in tumours [34-36].

The role of radiomics as independent predictors for identification of tumour tissue and characterising the signal patterns is evolving in pancreas cancer. The field has examined issues of diagnosis to separate benign from malignant pancreatic masses and patterns at base line to predict outcomes of surgical resection. Ours is the first study to correlate baseline signals with outcomes in the neoadjuvant setting. Clinical predictors of benefit from treatment have been examined for metastatic disease and genomic signatures that predict for outcome are also being researched. Previous studies of CT based prognostics that utilised traditional pathological parameters did make promising findings [37, 38], however our results suggest that there is a predictive radiomic pattern that should be tested in larger data sets. Improving selection of patients will improve cure rates and allow for more personalised therapy.

Relatively few investigations of the use of CT radiomic features for pancreatic cancer prognostics have considered multivariate models. A recent review carried out by He et. al in 2020 [39] described only two, Cheng et. al 2019 [40] who achieved an ROC AUC of 0.756 for predicting progression free survival in patients with unresectable PDAC receiving chemotherapy and Attiyeh et. al 2018 [41]who achieved a concordance index (a generalisation of ROC AUC) of 0.74 for predicting OS in chemotherapy-naive pre-surgical pancreatic ductal adenocarcinoma patients.

In a more comparable populations Koay et. al [42] investigated CT images of the interface between PDAC tumours and surrounding parenchyma in pretherapy PDAC patients and found that the difference in mean value of the Hounsfield unit (HU; a measure of radio density) distribution was predictive of distant metastasis free survival and overall survival in patients receiving neoadjuvant therapy (gemcitabine-based chemoradiation) or upfront surgery. Comparison of Kaplan-Meier curves do not indicate as great bifurcation as achieved by our multivariate model and multivariate assessments, including the radiomic feature, adjuvant chemotherapy and gender had concordance indices of 0.666 and 0.552 for T3 N0 and T3 N1 patients, respectively. In a follow up study [43] they demonstrated that a quantitative machine learning approach to the identification of this feature was possible and gave a highly predictive feature with hazard ratios of 2 and 1.9 for in multivariate models (including the single radiomic feature with patient, treatment and disease characteristics) predicting OS in patients receiving upfront surgery and gemcitabine-based chemoradiation, respectively. In another study Park et. al 2021 [12] predicted survival time after surgically resected PDAC from demographic, clinical and CT radiomic features, and achieved an improvement in the concordance index for the stratification of high-risk (survival <1 year) from low-risk (survival >3 years) patients from 0.6785 to 0.7414. The importance of the inclusion of patient and demographic factors along with radiomic features in prognostic modelling is highlighted by Permuth et. al 2021 [44] who investigated several CT radiomic features in a diverse population of patients who underwent pre-treatment imaging for PDAC. They demonstrated that some features’ correlation with survival was influenced by the ethnicity of the patient (in particular, African Americans with low volumetric mean HU tumours had worse survival than non-African Americans with the same tumour characteristics). Our work stands out for its precision in identifying a patient cohort who all have very short disease-free survival times (Fig. 4a) as well as a cohort who all have relatively long-term overall survival (Fig. 4b).

A limitation of this study is the small cohort size, which is a common issue in the medical imaging field due to the lack of image standardization between hospitals and institutions in addition to an absence of overarching registries or radiology image banks allowing the pooling of patients [45]. The scope defined for this study was to demonstrate the existence of radiomic signature sensitivity to tumour, treatment and also to the outcome. We have achieved this, however this project is not yet translational and these results should not be viewed as currently supporting implementation. Our findings are robust and hypothesis generating, however they require further validation in larger retrospective cohorts as well as in translational substudies in prospective trials.

## Supporting information

supplementary materials in one file

## Data Availability

All data produced in the present study are available upon reasonable request to the authors

