## supplementary materials in one file for "Computed Tomography Radiomics Signatures: Sensitive biomarkers for clinical decision support in pancreatic cancer- a pilot study"

### Supplementary table 1. Patient characterization

| PID | Inclusion/Exclusion | Reason for exclusion | age | cea | ca19 | bmi |
|---|---|---|---|---|---|---|
| GAP0001 | Excluded | PET Scan | 64.7 | 4.2 | 5 | 28.8 |
| GAP0002 | Excluded | PET Scan | 62.0 | 2.7 | 66 | 30.3 |
| GAP0003 | Included | NA | 63.4 | 4 | 70 | 24.4 |
| GAP0004 | Included | NA | 66.6 | 5.5 | 5 | 23.4 |
| GAP0005 | Excluded | PET Scan | 60.3 | 2.6 | 30 | 23.7 |
| GAP0006 | Included | NA | 64.5 | 2.1 | 13 | 34.0 |
| GAP0007 | Included | NA | 63.2 | 5.8 | 44 | 36.7 |
| GAP0008 | Excluded | Arterial phase only | 78.1 | 26 | 5441 | 21.1 |
| GAP0009 | Included | NA | 70.1 | 3 | 740 | 37.1 |
| GAP0010 | Included | NA | 49.1 | 2 | 221 | 30.4 |
| GAP0011 | Excluded | PET Scan | 59.9 | 1.6 | 1 | 29.9 |
| GAP0012 | Included | NA | 68.1 | 2 | 19 | 28.9 |
| GAP0013 | Included | NA | 53.6 | 25 | 296 | 26.2 |
| GAP0014 | Excluded | Technical problem | 65.9 | 6.2 | 360 | 29.0 |
| GAP0015 | Included | NA | 47.7 | 31 | 81 | 25.4 |
| GAP0016 | Included | NA | 54.3 | 5 | 1 | 25.2 |
| GAP0017 | Included | NA | 47.8 | 32 | 4359 | 28.7 |

| | | | | | | |
|---|---|---|---|---|---|---|
| GAP0018 | Included | NA | 71.0 | 2.7 | 1109 | 23.5 |
| GAP0019 | Included | NA | 66.8 | 2.5 | 3300 | 21.6 |
| GAP0020 | Included | NA | 66.3 | 0.7 | 133 | 26.6 |
| GAP0021 | Included | NA | 50.2 | 1 | 21 | 20.4 |
| GAP0022 | Excluded | PET Scan | 67.0 | 6.1 | 128 | 34.4 |
| GAP0023 | Included | NA | 77.6 | 5.3 | 440 | 25.4 |
| GAP0024 | Included | NA | 71.1 | 0.5 | 58 | 24.2 |
| GAP0025 | Included | NA | 58.3 | 3 | 2 | 19.0 |
| GAP0026 | Excluded | Empty folder | 69.7 | 1 | 37 | 25.4 |
| GAP0027 | Excluded | MRI Scan | 66.7 | 1.3 | 86 | 47.4 |
| GAP0028 | Excluded | Technical problem | 76.4 | 0.6 | 7 | 18.5 |
| GAP0029 | Included | NA | 68.3 | 3 | 470 | 21.3 |
| GAP0030 | Included | NA | 76.1 | 2 | 7305 | 26.4 |
| GAP0031 | Included | NA | 59.3 | 3 | 16 | 37.1 |
| GAP0032 | Included | NA | 62.4 | 1 | 80 | 34.1 |
| GAP0033 | Included | NA | 78.6 | 5.2 | 65 | 21.6 |
| GAP0034 | Included | NA | 66.8 | 1.3 | 562 | 27.0 |
| GAP0035 | Included | NA | 42.8 | | 47 | 19.5 |
| GAP0036 | Included | Technical problem | 67.6 | 6.5 | 1200 | 26.0 |
| GAP0037 | Excluded | PET Scan | 60.7 | 4.8 | 954 | 27.4 |
| GAP0038 | Excluded | PET Scan | 69.1 | 2.3 | 47 | 25.7 |
| GAP0039 | Included | NA | 75.7 | 5 | 21 | 25.0 |
| GAP0040 | Excluded | PET Scan | 63.7 | 1.3 | 1279 | 30.4 |
| GAP0041 | Excluded | PET Scan | 60.3 | 1.4 | 116 | 27.0 |
| GAP0042 | Included | NA | 58.0 | 1 | 274 | 20.7 |

**Supplementary figure 1. Consort flow diagram**

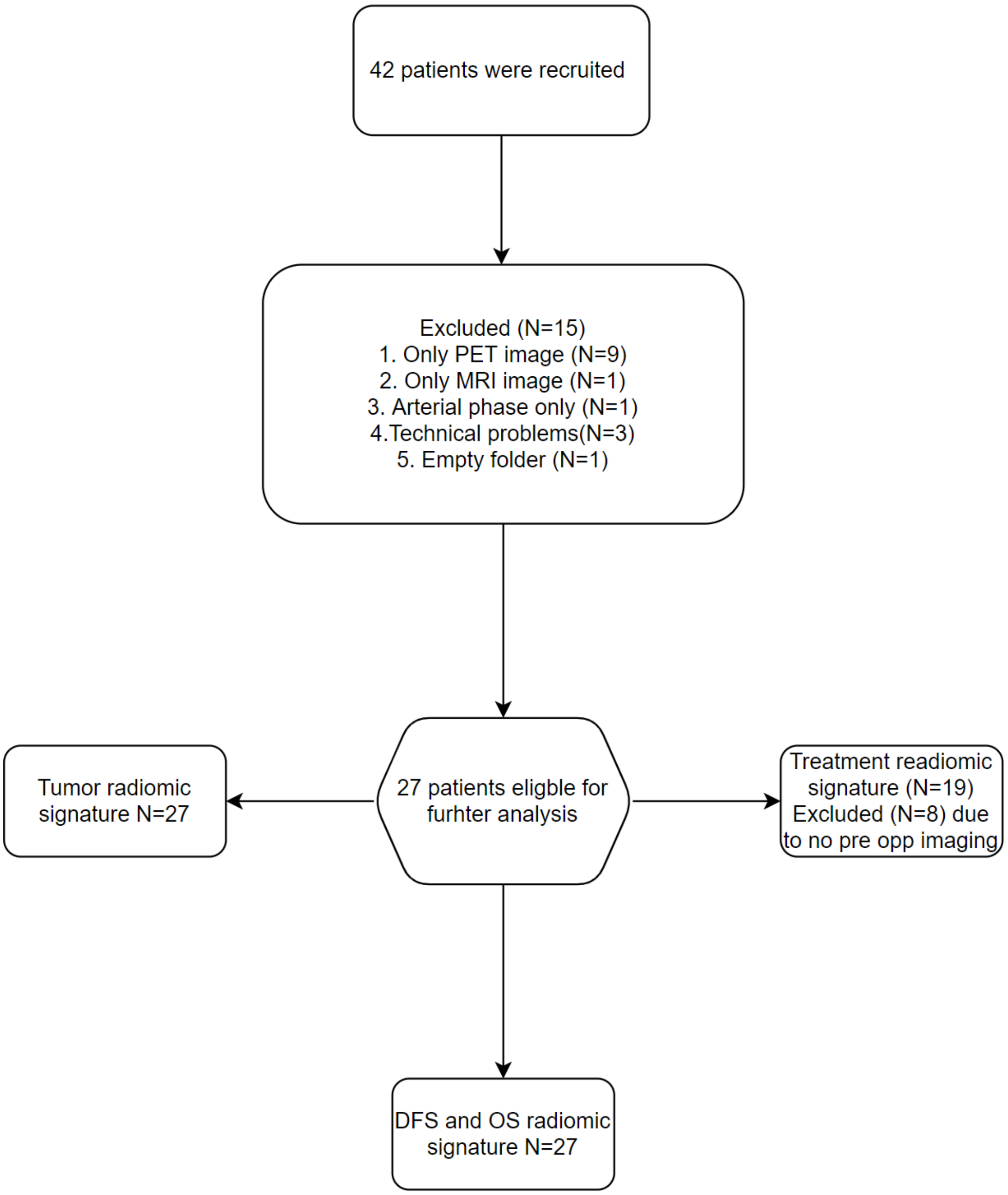


*Supplementary figure 1. Consort flow diagram representing the number of patients used in each radiomic signature.*

#### Supplementary Table 2. Tumor radiomic signature

| Feature No | Feature name | Coefficient (PC1) |
|---|---|---|
| 1 | DependenceNonUniformityNormalized | -0.21 |
| 2 | DependenceVariance | 0.19 |
| 3 | LargeDependenceEmphasis | 0.19 |

| | | |
|---|---|---|
| 4 | DependenceEntropy | 0.22 |
| 5 | SmallDependenceEmphasis | -0.15 |
| 6 | ZonePercentage | -0.14 |
| 7 | GrayLevelNonUniformity | 0.21 |
| 8 | SmallDependenceLowGrayLevelEmphasis | -0.20 |
| 9 | GrayLevelNonUniformity | 0.20 |
| 10 | ZoneVariance | 0.19 |
| 11 | LargeAreaLowGrayLevelEmphasis | 0.15 |
| 12 | LargeAreaEmphasis | 0.19 |
| 13 | ZoneEntropy | 0.19 |
| 14 | LargeDependenceHighGrayLevelEmphasis | 0.19 |
| 15 | Idmn | 0.17 |
| 16 | Busyness | 0.13 |
| 17 | Coarseness | -0.15 |
| 18 | RunLengthNonUniformity | 0.18 |
| 19 | Idn | 0.14 |
| 20 | DependenceNonUniformity | 0.18 |
| 21 | ShortRunLowGrayLevelEmphasis | -0.18 |
| 22 | TotalEnergy | 0.13 |
| 23 | LowGrayLevelRunEmphasis | -0.16 |
| 24 | LowGrayLevelEmphasis | -0.16 |
| 25 | GrayLevelNonUniformity | 0.16 |
| 26 | LargeAreaHighGrayLevelEmphasis | 0.17 |
| 27 | Median | -0.11 |
| 28 | Minimum | -0.17 |
| 29 | 90Percentile | -0.09 |
| 30 | LowGrayLevelZoneEmphasis | -0.15 |
| 31 | Mean | -0.11 |
| 32 | Contrast | -0.09 |
| 33 | GrayLevelNonUniformityNormalized | -0.15 |
| 34 | SmallAreaLowGrayLevelEmphasis | -0.14 |

### Supplementary Table 3. Treatment radiomic signature

| Feature no | Feature name | Coefficient (PC1) |
|---|---|---|
| 1 | DependenceNonUniformityNormalized" | -0.21 |
| 2 | "DependenceEntropy" | 0.22 |
| 3 | "Flatness" | 0.20 |
| 4 | "DependenceVariance" | 0.17 |
| 5 | "Coarseness" | -0.20 |
| 6 | "LeastAxisLength" | 0.21 |
| 7 | "Maximum2DDiameterColumn" | 0.20 |
| 8 | "LargeDependenceEmphasis" | 0.16 |
| 9 | "SmallDependenceLowGrayLevelEmphasis" | -0.20 |
| 10 | "ZoneEntropy" | 0.20 |
| 11 | "RunLengthNonUniformityNormalized" | 0.16 |

| | | |
|---|---|---|
| 12 | "SurfaceVolumeRatio" | -0.19 |
| 13 | "Maximum2DDiameterRow" | 0.18 |
| 14 | "Idmn" | 0.16 |
| 15 | "Maximum3DDiameter" | 0.18 |
| 16 | "ShortRunEmphasis" | 0.16 |
| 17 | "RunPercentage" | 0.16 |
| 18 | "Idn" | 0.12 |
| 19 | "SmallDependenceEmphasis" | -0.10 |
| 20 | "GrayLevelNonUniformityNormalized" | -0.17 |
| 21 | "LargeDependenceHighGrayLevelEmphasis" | 0.18 |
| 22 | "LowGrayLevelRunEmphasis" | -0.19 |
| 23 | "ZonePercentage" | -0.10 |
| 24 | "MinorAxisLength" | 0.20 |
| 25 | "LowGrayLevelEmphasis" | -0.18 |
| 26 | "SurfaceArea" | 0.19 |
| 27 | "MajorAxisLength" | 0.14 |
| 28 | "LowGrayLevelZoneEmphasis" | -0.18 |
| 29 | "SmallAreaLowGrayLevelEmphasis" | -0.17 |
| 30 | "ShortRunLowGrayLevelEmphasis" | -0.18 |

### Supplementary Table 4. DFS radiomic signature

| Feature No | Feature name | Coefficient |
|---|---|---|
| 1 | ZoneEntropy | -1.19 |
| 2 | "GrayLevelNonUniformityNormalized" | -0.82 |
| 3 | "LeastAxisLength" | -0.85 |
| 4 | "Maximum2DDiameterRow" | 1.14 |
| 5 | "SmallDependenceLowGrayLevelEmphasis | -0.21 |

### Supplementary Table 5. OS radiomic signature

| Feature No | Feature name | Coefficient |
|---|---|---|
| 1 | ZoneEntropy | -0.99 |
| 2 | ClusterTendency | -9.05 |
| 3 | Correlation | 1.21 |
| 4 | DifferenceVariance | 8.40 |
| 5 | RootMeanSquared | 0.50 |

### Supplementary figure 2: DFS and OS classification

**Description:** The DFS and OS radiomic signatures were fed to a linear classifier to predict patient group labels. The ROC corresponding to DFS and OS was cross validated with AUC found to be 0.94±0.07 and 0.87±0.17, respectively.

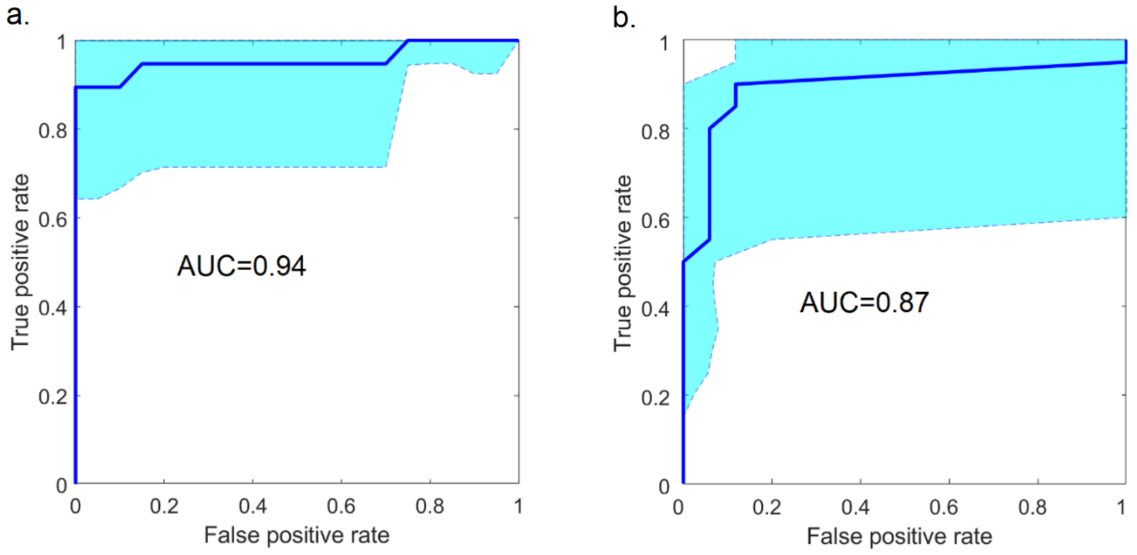


*Supplementary figure2. (a) ROC corresponding to DFS to classify long vs. short survival time with AUC=0.94±0.07. 95% confidence interval shaded in blue (b) ROC corresponding to OS to classify long vs. short survival time with AUC=0.87±0.17. 95% confidence interval shaded in blue.*
